# Retrospective Quality Analysis of a Clinical RAG Chatbot: Observable Signals and Lessons Learned

**DOI:** 10.64898/2026.01.26.26344757

**Authors:** Ilia Khashei, Damiano Presciani, Luca Paolo Martinelli, Simon Grosjean

## Abstract

Retrieval-augmented generation (RAG) is increasingly adopted to ground clinical conversational agents in external knowledge sources, yet many deployed prototypes lack the observability required for standard RAG evaluation. In particular, retrieved documents and grounding context are often not logged, preventing direct assessment of retrieval quality and faithfulness. We report a post-hoc evaluation of *EMSy*, a clinical RAG-based chatbot prototype, based on 2,660 multi-turn conversations collected between January and September 2025. Rather than benchmarking performance, we adopt an evaluation strategy based exclusively on observable signals. The analysis combines an exploratory intent analysis conducted on a random subset of heterogeneous interactions, automated quality scores available at the message and conversation level, and explicit user feedback, with 96.0% of rated conversations receiving positive feedback. Results indicate that message-level minimum scores capture localized low-quality responses that are not reflected by average conversation-level metrics, while user feedback reflects aggregate interaction impressions. This case study illustrates how diagnostic insights can be obtained under limited observability and identifies implications for the design and evaluation of future clinical RAG systems.

## 1 Introduction

Large language models are increasingly explored for clinical conversational applications, including decision support, information delivery, and triage-oriented systems. To mitigate hallucinations and improve factual grounding, many systems adopt retrieval-augmented generation (RAG), in which model outputs are conditioned on information retrieved from external knowledge sources. Despite their growing use, systematic evaluation of clinical RAG systems remains challenging, particularly outside controlled benchmark settings.

Recent evaluation frameworks for RAG systems rely on access to retrieved passages, relevance rankings, and grounded model inputs in order to assess retrieval quality, grounding faithfulness, and answer correctness. These assumptions frequently do not hold in early-stage or legacy clinical prototypes, where intermediate artifacts may not be logged due to architectural, operational, or privacy constraints. As a result, standard RAG evaluation methodologies are often inapplicable to real-world systems analyzed retrospectively.

In this work, we present a post-hoc evaluation of *EMSy*, a clinical RAG-based chatbot prototype, conducted under limited observability conditions. Retrieved documents, relevance scores, and grounded prompts were unavailable, precluding direct component-level evaluation. Rather than treating this setting solely as a limitation, we frame it as a common scenario in clinical AI development and examine which evaluation signals remain informative when only inputs, outputs, and aggregate signals are observable.

The objective of this study is methodological rather than comparative. We do not benchmark EMSy against alternative systems or claim clinical effectiveness. Instead, we investigate how interaction-level intent, automated quality scores, and user feedback can be combined to characterize system behavior, identify localized failures, and inform future system design under realistic observability constraints.

## 2 Related Work

Retrieval-augmented generation (RAG) has become a common architectural approach for grounding large language model outputs in external knowledge sources, particularly for knowledge-intensive tasks [3, 2]. By decoupling knowledge retrieval from generation, RAG systems aim to improve factual accuracy and reduce hallucinations, making them especially appealing in high-stakes domains such as healthcare.

Several evaluation frameworks have been proposed to assess RAG systems, focusing on retrieval quality, grounding faithfulness, and answer correctness with respect to retrieved evidence [1]. These approaches typically assume full observability of the retrieval process, including access to retrieved documents, relevance rankings, and grounded model inputs. While appropriate for controlled experimental settings, such assumptions often do not hold in early-stage or legacy systems, where intermediate artifacts may not be logged due to architectural, operational, or privacy constraints. In parallel, a growing body of work has examined the use of large language models in clinical and biomedical contexts, highlighting both their potential capabilities and their limitations [5, 7]. Prior research has emphasized the importance of transparency, auditability, and careful evaluation when developing and deploying clinical AI systems [4, 6]. However, comparatively less attention has been paid to methodological strategies for evaluating clinical conversational systems under partial observability constraints.

Recent surveys of conversational agents in healthcare summarize design patterns, evaluation challenges, and deployment considerations across clinical settings [9]. Recent work has also specifically examined the application of retrieval-augmented generation in medical contexts, discussing opportunities, challenges, and practical considerations for clinical deployment [8]. Our study complements this line of research by focusing on post-hoc evaluation strategies that rely exclusively on observable interaction-level signals when retrieval-level artifacts are unavailable.

## 3. System Description and Data

### 3.1 System Overview

*EMSy* is a prototype clinical conversational agent designed to support queries related to emergency medicine and clinical decision making. The system follows a retrieval-augmented generation architecture, in which user inputs trigger retrieval from a curated clinical knowledge base, and a large language model generates responses conditioned on the retrieved content. The system was developed for exploratory and research purposes rather than deployment in clinical care.

The chatbot supports multi-turn conversations. System responses are intended to be informative and cautious and do not provide diagnoses or replace professional medical judgment. Details of the underlying language model, retrieval algorithms, and knowledge sources are not required for the methodological focus of this study and are therefore omitted.

### 3.2 Interaction Logs

The analysis is based on retrospective interaction logs collected during use of the EMSy prototype. The dataset comprises 2,660 multi-turn conversations collected between January and September 2025. In total, the logs contain 46,355 messages, including 20,261 user messages and 26,094 system responses.

Automated quality scores are available for all system responses and are aggregated at the conversation level. Explicit user feedback in the form of upvotes or downvotes is available for 199 conversations, corresponding to 7.5% of the dataset.

At conversation start, users were asked to self-identify their professional role. Available role labels include physician (698 conversations), nurse (532), emergency medical technician (77), and unspecified (1,327). Role information was not used to condition system behavior and is reported solely for descriptive purposes.

Conversations are also associated with coarse system-level categories reflecting internal routing within the prototype. Among these, a residual category labeled “varie/altro” aggregates interactions that do not fall into predefined structured categories. This category contains heterogeneous, free-form interactions and is treated in this study as a dataset attribute without intrinsic semantic interpretation.

No personally identifiable information was included in the analyzed data. All analyses were conducted retrospectively for methodological evaluation.

### 3.3 Observability Constraints

The interaction logs do not include retrieval-level artifacts. Retrieved documents, relevance scores, grounded prompts, and citation information are unavailable. Consequently, direct evaluation of retrieval quality, grounding faithfulness, and evidence attribution is not possible, and standard RAG evaluation frameworks cannot be applied. All analyses presented in this paper are explicitly designed to operate under these observability constraints.

## 4 Evaluation Setting Under Limited Observability

Under limited observability, evaluation is restricted to system inputs and outputs. For each conversation, observable data consist of user messages, system responses, automated quality scores, and optional user feedback. The retrieval process and grounded model inputs remain latent.

In this setting, evaluation shifts from component-level validation to behavioral characterization. Rather than assessing whether correct evidence was retrieved or faithfully used, the analysis focuses on observable response patterns, score distributions, and relationships between proxy signals. This perspective reflects common conditions in retrospective analyses of early clinical AI systems.

## 5 Methods

### 5.1 Evaluation Strategy

We adopt a descriptive evaluation strategy based exclusively on observable signals. The analysis integrates exploratory intent annotation within a heterogeneous subset of conversations, automated quality scores at the message and conversation level, and explicit user feedback. No ground-truth correctness labels are assumed.

### 5.2 Exploratory Intent Analysis

Intent analysis is restricted to conversations labeled as “varie/altro”, comprising 500 conversations. From this subset, a random sample of 150 conversations was selected for intent annotation.

Each sampled conversation was assigned a single dominant intent label based on the overall user interaction. Five coarse-grained intent categories were defined: pharmacological or therapeutic queries, procedural queries, educational or explanatory requests, clinical decision or assessment queries, and meta-level interactions. Intent labels were used for descriptive stratification and were not treated as causal explanations of system behavior.

### 5.3 Automated Quality Scores

Automated quality scores are available for all system responses. Scores are defined on a continuous scale from 0 to 1 and are treated as relative internal signals rather than validated measures of clinical quality or safety. For each conversation, both the mean score across system responses and the minimum score observed within the conversation are analyzed, with the latter used as a proxy for worst-case behavior.

### 5.4 User Feedback

User feedback is provided at the conversation level in the form of upvotes or downvotes. Feedback is optional and reflects users’ overall subjective assessment of the interaction. Feedback signals are analyzed descriptively and in relation to automated scores.

## 6 Results

### 6.1 Dataset Overview

All 2,660 conversations contain at least one system response with an associated automated quality score. User feedback is available for 199 conversations (7.5%). No external correctness annotations or expert labels are available.

### 6.2 Intent Distribution in the “varie/altro” Subset

Figure 1 reports intent proportions estimated from the random sample of 150 “varie/altro” conversations. Pharmacological or therapeutic queries account for 34.0% of sampled conversations, followed by procedural queries (24.7%), educational or explanatory requests (18.0%), and clinical decision or assessment queries (16.7%). Meta-level interactions account for 4.7%, while administrative and guideline-related intents together account for less than 2%. These estimates characterize interaction diversity within the heterogeneous subset and are not intended to represent the full dataset.

**Figure 1.**
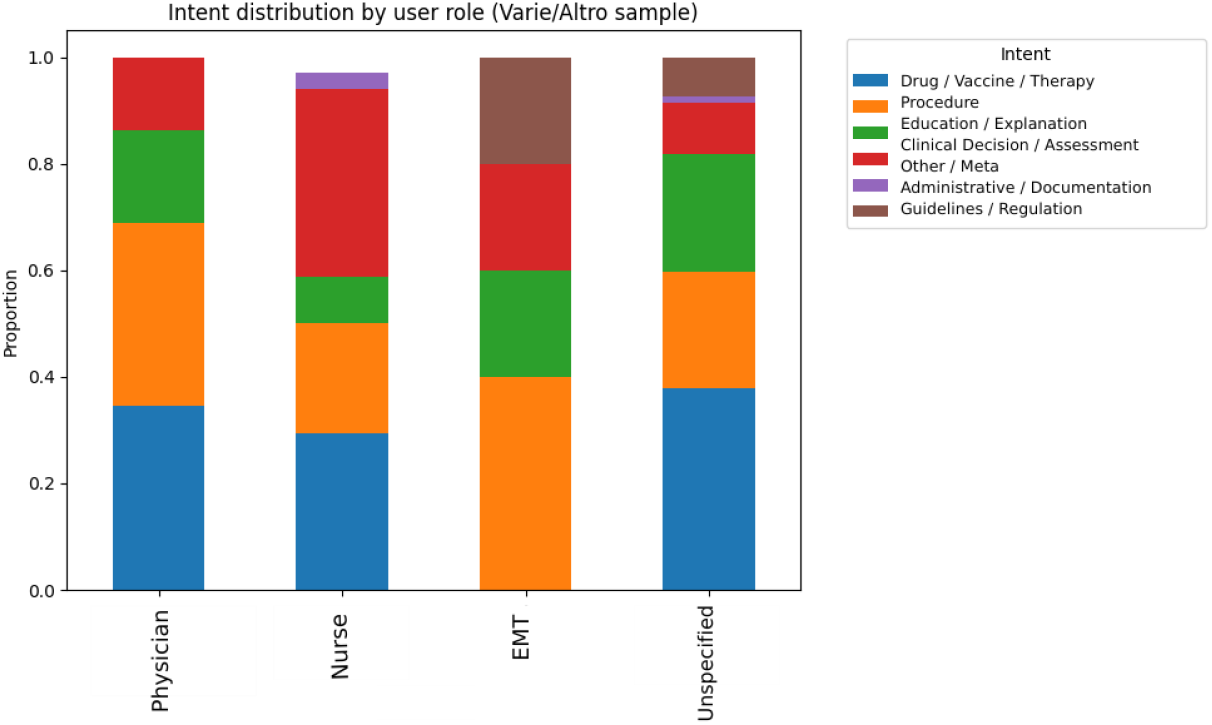
Estimated distribution of dominant user intents in a random sample of 150 conversations from the “varie/altro” subset.

### 6.3 Automated Quality Scores

Conversation-level mean automated scores exhibit substantial variability across interactions. Message-level scores are skewed toward higher values but display a long tail of lower scores, indicating the presence of localized low-quality responses.

Across conversations, minimum message-level scores are consistently lower and more variable than mean scores. This indicates that conversations with relatively high average quality may nonetheless contain individual responses with markedly lower scores. Within the “varie/altro” subset, differences in minimum score distributions are observed across intent categories, with scenario-based and evaluative intents exhibiting lower minima than purely informational queries.

### 6.4 User Feedback and Automated Scores

Among the 199 conversations with feedback, 191 (96.0%) received positive feedback and 8 (4.0%) received negative feedback. This distribution indicates overall favorable user reception within the subset where feedback was provided.

Conversations receiving negative feedback tend to exhibit lower automated scores on average than positively rated conversations, although score distributions overlap. Differences between upvoted and downvoted conversations are more pronounced when considering minimum message-level scores rather than mean scores. Downvoted conversations are more likely to include at least one low-scoring response, even when average scores remain moderate.

## 7 Discussion

This study presents a methodological analysis of post-hoc evaluation for a clinical RAG system under limited observability. By combining intent stratification, automated quality scores, and user feedback, we characterize system behavior using signals that remain accessible in the absence of retrieval-level evidence.

The results indicate that message-level minimum scores capture localized low-quality responses that are not reflected by conversation-level averages and align more closely with negative user feedback. This suggests that worst-case aggregation provides complementary diagnostic value in settings where safety considerations are salient and ground-truth labels are unavailable.

### 7.1 System Evolution and Future Evaluation

The analyses reported here are based on interaction logs from an early EMSy prototype employing a simple RAG architecture with limited logging. Retrieval-level artifacts were not stored, constraining the scope of retrospective evaluation. Since the period covered by this study, EMSy has been redesigned and is now operational with a revised architecture that prioritizes auditability and transparency. The current system supports inspection of retrieval behavior and grounding signals, enabling direct evaluation of RAG components. Future work will focus on evaluating the redesigned system and assessing the impact of improved instrumentation on evaluability.

## 8 Limitations

This study is limited by the absence of retrieval-level artifacts, which prevents direct assessment of retrieval quality and grounding faithfulness. Intent analysis is restricted to a random sample of conversations within a heterogeneous subset and is not representative of the full dataset. Automated quality scores are treated as relative internal signals rather than validated measures of clinical performance. User feedback is optional and available for a limited subset of conversations, which constrains its interpretability.

## 9 Conclusion

We presented a post-hoc observational evaluation of a clinical RAG chatbot conducted under limited observability. Using only observable signals, we demonstrated how localized failures and interaction patterns can be identified without access to retrieval artifacts. The findings emphasize the importance of message-level analysis and worst-case metrics in clinical conversational systems and highlight the need for future RAG architectures to support auditability and systematic evaluation by design.

## Data Availability

The data analyzed in this study consist of retrospective, anonymized interaction logs from a proprietary clinical chatbot system (EMSy). Due to privacy, ethical, and commercial constraints, the raw interaction data are not publicly available. Aggregated data and summary statistics supporting the findings are included in the manuscript. Further details may be provided by the authors upon reasonable request, subject to applicable restrictions.

## Acknowledgments

The authors would like to thank the entire EMSy team for their support in the development and maintenance of the platform. In particular, we extend our gratitude to Nicolò Balzani for his strategic vision and support during the data collection phase. We also thank the emergency professionals who participated in the testing phase and provided valuable feedback that informed this analysis.

## Conflict of Interest

S.G. is the CEO and founder of EMSy. D.P. is the CTO and co-founder of EMSy. L.M. is a member of the founding team. The system analyzed in this study (EMSy) is a proprietary product developed by the authors. To mitigate potential bias, the evaluation was conducted using retrospective, anonymized interaction logs and standard automated metrics. The authors report no additional competing financial interests beyond those disclosed above.

## Notes

### Funding Statement

This study did not receive any external funding.

### Author Declarations

Ethics committee of EMSy S.r.l. waived ethical approval for this work. The study involved retrospective analysis of fully anonymized interaction logs collected during routine system use and did not involve any intervention, patient contact, or collection of identifiable personal data.

